# Precise dose verification in proton therapy using Positron Emission Tomography

**DOI:** 10.1101/2025.08.02.25332864

**Authors:** Marcin Balcerzyk, Marta Freire, Rubén Fernandez de la Rosa, Miguel Angel Pozo, Rhodri Lyn Smith, Gaspar Sanchez-Merino, Antonio Gonzalez

## Abstract

**Background:** Proton and ion therapy have gained significant importance in radiation therapy cancer treatment due to their favorable dose distribution and tissue-sparing properties. In conventional gamma radiation therapy some methods of *in vivo* dose verification are possible with current medical devices. Proton and ion therapy dose verification is limited, mainly using PET for particle range. Prompt gamma methods offer low spatial resolution. This study presents initial results for in-vivo dose verification with PET imaging of F-18 during proton therapy. Although the activity concentration of F-18 generated by typical clinical doses (several Gy) is low, PET imaging performed approximately one hour post-irradiation yields sufficient image quality to derive dose-volume histograms (DVH), enabling spatial dose verification.

**Purpose:** To verify the applied dose in proton therapy *in vivo* using Positron Emission Tomography with millimetric precision.

**Materials and Methods:** We simulated proton treatment in a brain phantom using Gate and RayStation platforms to assess the production of several positron emitting isotopes. We focused on the production of fluorine-18 (F-18), given its low positron energy, which enables accurate reproduction of the dose distribution. To evaluate the detectability of the anticipated low activity concentrations (on the order of a few Bq/mL) following a 3 Gy proton irradiation, we tested three PET systems: two preclinical scanners based on LYSO detectors and one clinical scanner based on BGO crystals. Finally, we have analyzed the dose-volume histograms for simulated and measured dose and activity distributions and compared them with the planned ones.

**Results:** F-18 PET imaging in proton therapy correlates with delivered dose within 5% error and matches the planned dose fall-off edge within 1 mm, enabling accurate and precise *in vivo* dose verification.

**Conclusion:** The dose verification in proton therapy using F-18 Positron Emission Tomography allows higher precision of dose than other positron emitters like C-11, N-12 or O-15.

**Key Results:** **Key Results:** In proton therapy, in-vivo F-18 production correlates with the deposited dose in the patient with a 5% margin of error. The leading-edge position of the F-18 activity distribution agrees with the planned dose fall off within 1 mm. Furthermore, the feasibility of detecting low activity concentrations typical of proton therapy (Bq/ml range) has been demonstrated using both preclinical and clinical PET scanners.

**Required Summary Statement:** In proton therapy the delivered dose to the patient can be measured *in vivo* using Positron Emission Tomography by imaging the production of F-18 isotope, achieving millimetric spatial accuracy.

## 3 Introduction

Proton and other hadron therapies are reaching a mature stage in radiation treatment of cancer. There are more than 100 centers in the world with several being currently under construction. A critical component of radiation therapy is the verification of the radiation treatment plan. Currently, in both gamma and proton therapies, verification is performed prior to patient irradiation using devices such as wire chamber arrays which offer spatial resolutions on the order of several millimeters. However, this pre-treatment verification does not guarantee that dose is actually delivered as planned during therapy. Failure to verify actual dose delivery can result in inadequate cancer treatment or damage to surrounding healthy tissues. This is exemplified by incidents in France between 2003-2007, where over 800 patients received incorrect doses in gamma radiotherapy, leading to severe complications(1). To avoid such risks, it is crucial to verify the dose delivered to the patient during or after each treatment session, not only at the planning stage.

In gamma radiotherapy, in-vivo dose verification is already being implemented in emerging commercial systems. One such example is the VeriQA system from PTW Dosimetry (Germany)(2). In this approach, a 6 MV treatment beam from a 6 MV accelerator, after passing through a multi-leaf collimator, is captured after traversing the patient. From this captured radiation, the actual dose absorption that has occurred in the patient is reconstructed and compared with the expected dose according to the treatment planning CT. This allows for the identification of discrepancies between the planned and administered treatment. While the spatial resolution and accuracy of dose verification depend on multiple circumstances, the final result is suitable for detecting errors in treatment administration.

This approach is not feasible in proton or hadron therapy, as the particles are fully stopped in the patient’s body and they do not exit the tissue to be detected externally. Therefore, several methods have been developed for dose verification specifically for these modalities.

1. **Prompt Gamma Detection** When protons interact with atomic nuclei in the patients tissues they can induce the emission of prompt gamma rays. These gamma rays can be detected in real time and provide an indirect measure of proton range. However, the accuracy of this method is limited to about 5 millimeters, which is less precise than the 1-millimeter accuracy desired by radiation physicists. Detecting these gamma rays in the electrically noisy environment of a treatment room presents significant challenges (3-5)
2. **Positron Emission Tomography (PET)** This approach involves detecting positron-emitting isotopes produced when protons interact with certain elements in the body, such as oxygen, nitrogen, and carbon(6). PET imaging provides quantitative, three-dimensional information on the spatial distribution of these isotopes in the 1-2 mm range, offering insight into the location and extent of proton interactions within the body. However, the isotopes used in this method, such as O-15, N-13, N-12 and C-11, have relatively short half-lives, making it difficult to accurately measure the dose during or immediately after the treatment session(7-10). Further complicating the interpretation, these isotopes often form gaseous species that react with oxygen and diffuse rapidly through tissue, blurring the spatial correlation between the site of production and the PET signal. Additionally, the heterogeneous elemental composition of different tissues alters isotope production rates, introducing further uncertainty into dose mapping (11, 12).
3. **Use of Contrast Agents** Medical physicists sometimes use contrast agents to enhance the visibility of proton interactions within the body. For instance, injecting in the tumor area water enriched with the isotope O-18 can help produce F-18, an isotope that emits positrons and can be measured using PET. However, this approach requires the injection of additional substances into the body, adding complexity to the treatment(11, 12).

The methods above are helpful but have limitations in terms of accuracy, practicality and interpretation. The complex nature of the nuclear reactions involved, as well as the variability in tissue composition, makes it difficult to precisely determine the proton and more importantly, they often fall short in verifying the actual dose delivered to the tumor. Importantly, range verification alone is insufficient—what is ultimately required is direct, spatially resolved dose verification.

To address this, we propose a novel strategy: using the well-known nuclear reaction ^18^O(p,X)^18^F as the basis for dose verification in proton therapy. This reaction occurs naturally due to the presence of ^18^O in tissue water, albeit at low natural abundance (∼0.2%). This reaction is well-known with a moderate cross-section (500 mb maximum, c.f. Suppl. Fig. 3 A) and used in commercial production of F-18 radiotracers for PET(13). It has a low threshold of about 2 MeV (Suppl. Fig. 3 A). In this paper we examine through simulations the production of β+ isotopes derived from these reactions in phantoms of brain tissue, we present the results and feasibility of low activity concentrations measurements in PET scanners and finally we examine the dose-volume histograms from the simulations and measurements that use F-18. This method of dose verification using β+ isotopes in proton and He-4 radiation therapy is patent pending (EP25382365).

## 4 Materials and Methods

### 4.1 Simulations with Gate

For simulations of proton and alpha beam interactions with tissues we used the Gate program with version 10.0 (14) and 9.3(15). Due to high interest in treatment of brain tumors we used brain tissue that has the composition defined in GateMaterials.db(16). The composition is shown in Table 1 and recalculated to molar concentration and fraction. Gate recommends the use of physics lists of processes. For the simulations we have used QGSP_BERT_HP_EMY, which is recommended for collider physics applications with electromagnetic components as option 3(17).

**Table 1.** Composition of brain tissue according to(16).

| Brain density 1.04 g/cm<sup>3</sup> | Molar mass, g | mass fraction | g in 1L | mol in L | mol fraction |
| --- | --- | --- | --- | --- | --- |
| Hydrogen | 1.01 | 0.107 | 111.28 | 110.1782 | 0.64383 |
| Carbon | 12.01 | 0.145 | 150.8 | 12.5562 | 0.07337 |
| Nitrogen | 14.01 | 0.022 | 22.88 | 1.633119 | 0.00954 |
| Oxygen | 16 | 0.712 | 740.48 | 46.28 | 0.27044 |
| Sodium | 22.99 | 0.002 | 2.08 | 0.090474 | 0.00053 |
| Phosphor | 30.97 | 0.004 | 4.16 | 0.134324 | 0.00078 |
| Sulfur | 32.066 | 0.002 | 2.08 | 0.064866 | 0.00038 |
| Chlorine | 35.45 | 0.003 | 3.12 | 0.088011 | 0.00051 |
| Potassium | 39.098 | 0.003 | 3.12 | 0.079799 | 0.00047 |
| Calcium | 40.08 | 0 | 0 | 0 | 0 |
| Scandium | 44.956 | 0 | 0 | 0 | 0 |

We first simulated a monoenergetic 75 MeV proton beam (1 mm FWHM, parallel geometry) directed at the brain phantom. We tracked the deposited dose from 10^6^ protons and quantified the production of β^+^-emitting isotopes: N-12, N-13, O-15, and F-18 (see Suppl. Fig. 1 for dose distribution). The cross sections for these reactions with protons are shown in Suppl. Fig. 3. The macro files for Gate 9.2 are provided in Supplemental Material 1 zip file.

**Figure 1.**
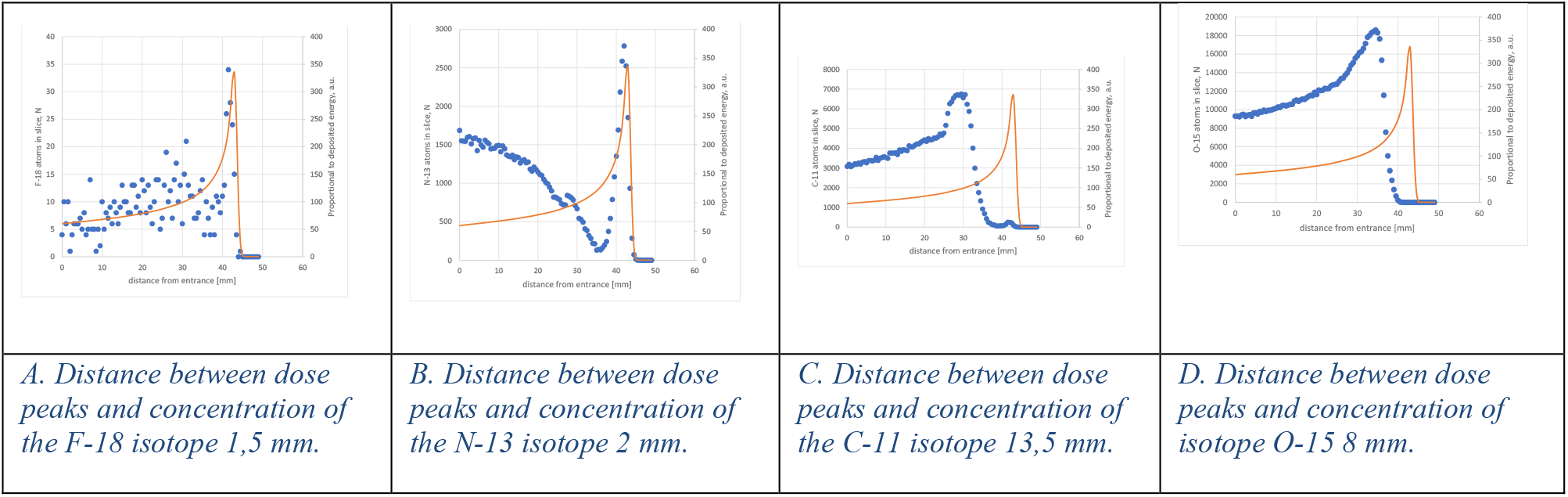
Simulations. A value proportional to the deposited dose (orange line) and the number of positron-emitting isotope atoms produced per slice (blue dots) versus distance from the entrance with 75 MeV proton beam.

We then simulated a treatment with proton beam of a 3×3×4 cm^3^ tumor at the depth of 8.4 cm of its center in a brain tissue. The cubic form was used to easily analyze leading edge positions, difference between deposited dose and distribution of β+ isotopes. The pencil beam nozzle was located 50 cm from the isocenter. The energy spread was 1%. The required six energies were from 93 to 123 MeV for protons. The matrix of points was 6×6 mm in a plane perpendicular to the beam. The data for x y theta phi and emittance are provided in the macro files for Gate 9.2 in Supplemental Material 2 zip file. We simulated 5 ×10^9^ protons using VIP cluster in vip.creatis.insa-lyon.fr(18).

All voxel-based images were generated with 1 mm pixel resolution. Our aim was to analyze the true spatial distribution of β^+^-emitting isotopes (not PET reconstructions) for comparison with the planned dose distribution. The degradation introduced by PET image reconstruction, scanner geometry, calibration, and acquisition time is addressed in the following section through physical scanner measurements.

### 4.2 Low activity concentration measurements in PET scanners

Given the expected low activity concentration of F-18 from clinical doses, we evaluated the feasibility of detection using three PET systems:

The first was the Bruker Albira preclinical PET/CT system described in(19). The used system version has two detector rings and 5% peak sensitivity and 5 ns coincidence window. The detectors are based on LYSO crystals (5% yttrium, 95% lutetium). We performed a 12 h study without decay correction of 3 mL syringe with starting [F-18]FDG activity concentration of 1 kBq/mL which decayed to 11 Bq/mL. The study was reconstructed in 1 h windows, which was used for subsequent analysis.

The second scan was to verify that the source of the noise seen in the low activity concentration is not electronic. We used a custom-built scanner for PET mammography named DeepBreast(20). The detectors are also based on LYSO crystals (10% yttrium, 90% lutetium). It has 2% sensitivity and 3 ns coincidence window. We scanned a 20.25 mL [F-18]FDG phantom with an initial activity concentration of 200 kBq/mL. We performed a 33 h study without decay correction with final activity concentration of 0.73 Bq/mL. We chose the volume of 20 mL as this is the typical one for the tumors treated in radiation therapy.

To positively verify that the elimination of lutetium in the PET detector crystals removes the noise in the image, we used a BGO-based clinical PET scanner, namely GE Healthcare Omni Legend 32 described in(21, 22). We carried out a 24 h study without decay correction of 35 mL vial with starting activity concentration of 1 kBq/mL which decayed to 0.3 Bq/mL.

### 4.3 Dose-volume-histogram analysis

Using Gate simulations, we evaluated the production of C-11, N-12, N-13, O-15 and F-18 following the planned proton therapy treatment. We calculated the dose from the activity concentrations of these isotopes, as explained in the Results section. We focused on the planned treatment volume (PTV) and generated histograms representing the number of C-11 and F-18 atoms produced within this volume. To facilitate comparison with the planned dose distribution, we aligned the 50% dose value (D50) with the half-height of the C-11 and F-18 activity histograms. This allowed us to derive a conversion factor to map activity concentration (Bq/mL) to dose (Gy). The resulting Dose–Volume Histograms (DVHs) based on isotope production were then compared to those from the original RayStation treatment plan, assessing spatial concordance between the simulated biological response (β^+^ isotope distribution) and physical dose deposition.

## 5 Results

### 5.1 Simulations

We simulated the interaction of monoenergetic 75 MeV proton beams with brain-equivalent tissue to evaluate the spatial correlation between dose deposition and β^+^- emitting isotope production. The results are presented in Figure 1. It shows the distance between dose peak and β+ isotope distribution for F-18 (A), which is only 1.5 mm, for N-13 (B) which is 2 mm, but the reproducibility of the dose for entering protons shows high discrepancy. For C-11 (C) the distance between peaks is already 13.5 mm and for O-15 (D) it is 8 mm. For N-13, C-11 and O-15 t the dose for entering protons shows high discrepancy. However, for F-18, the dose shows a high similarity.

In Figure 2 we show the simulated 3D distribution of dose, C-11, and F-18 following irradiation of a cubic 3×3×4 cm^³^ tumor phantom with a peak dose of 0.74 Gy from 5×10^9^ protons. The upper row presents the dose distribution; the middle and lower rows show the corresponding C-11 and F-18 isotope distributions, respectively.

**Figure 2.**
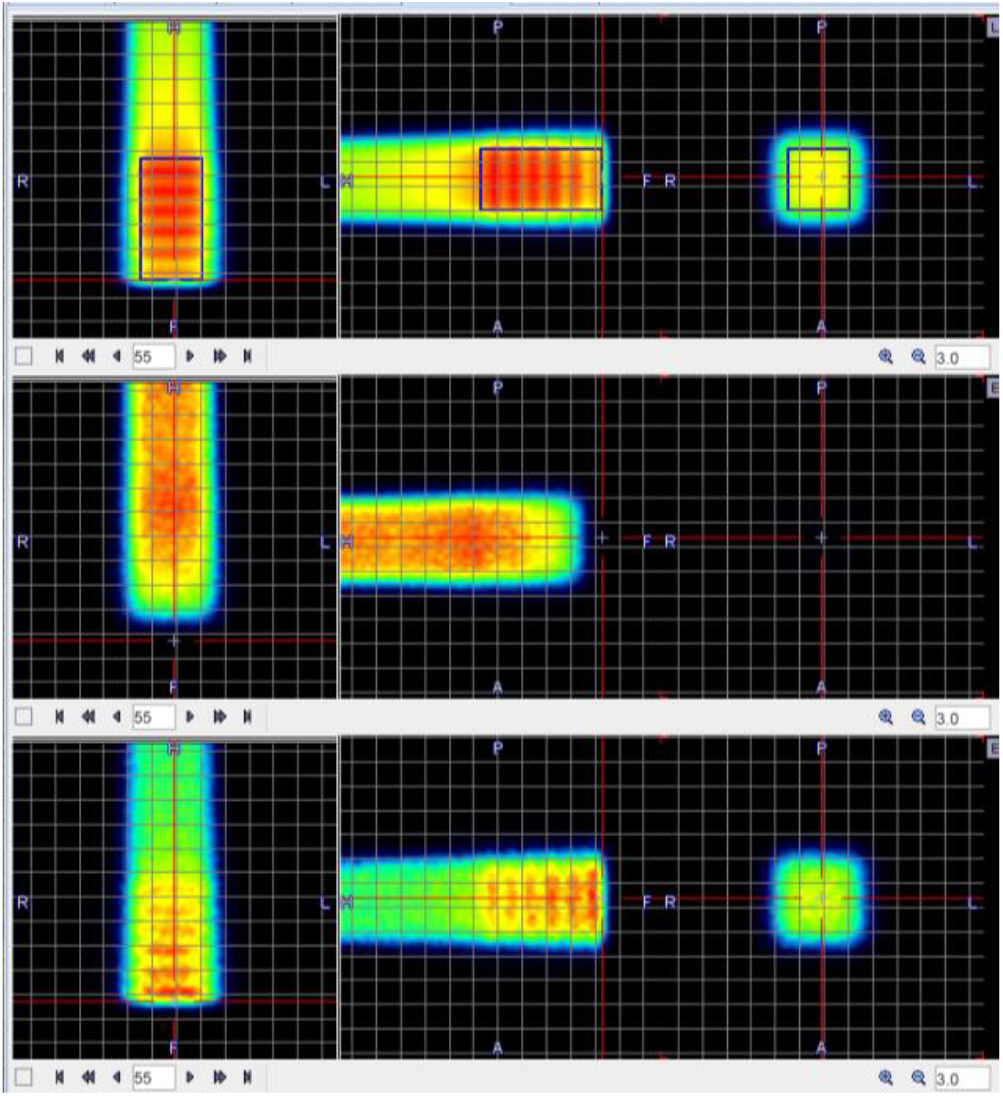
Simulation. **Top**: dose (Gy) in 3×3×4 cm cube with 5×10^9^ protons reaching 0.74 Gy. **Middle**: C-11 initial activity map with 0.7 Gy dose reaching 43 Bq/mL at therapy start. **Bottom**: F-18 initial activity map with 0.7 Gy dose reaching 0.022 Bq/mL at therapy start. The red cross cursor is positioned at the front of the irradiated cube, note that F-18 activity represents the dose very well, while C-11 is misplaced by about 25 mm without dose proportional representation. The grid is 10 mm.

The F-18 image closely reproduces the leading edge of the dose distribution, whereas the C-11 distribution is misplaced by ∼25 mm toward the beam origin.

The temporal evolution of β^+^ activity across the phantom (see Suppl. Fig. 2) reveals that O-15 dominates during the first 10 minutes post-irradiation, followed by C-11 up to 150 minutes, and finally F-18, due to its 110-minute half-life, dominates thereafter. These time courses do not account for the ∼10-minute treatment duration, which would further reduce early isotope contributions. Washout effects for C-11 and O-15 were modeled using physiological clearance data from rabbit tissue (11, 12), and conversion factors for activity-to-dose mapping accounting for washout are summarized in Table 2.

**Table 2.** Activity concentration to dose conversion factors for brain tissue for several times after treatment. The bottom part takes into account the washout of C-11 and O-15 from (12) using data for rabbit brain.

| Activity,<br>Bq/mL per 1<br>Gy for $\beta^+$ | t=0 min | t=10<br>min | t=30<br>min | t=60<br>min | t=150<br>min | t=180<br>min | t=210<br>min | t=240<br>min | t=270<br>min |
| --- | --- | --- | --- | --- | --- | --- | --- | --- | --- |
| O-15 | 1042 | 34.65 | 0.04 | 0.00 | 0.00 | 0.00 | 0.00 | 0.00 | 0.00 |
| N-13 | 29.71 | 14.82 | 3.69 | 0.46 | 0.01 | 0.00 | 0.00 | 0.00 | 0.00 |
| F-18 | 0.0317 | 0.0298 | 0.0262 | 0.0217 | 0.0149 | 0.0123 | 0.0102 | 0.0084 | 0.0070 |
| C-11 | 61.6724 | 43.9063 | 22.2536 | 8.0299 | 1.0455 | 0.3773 | 0.1361 | 0.0491 | 0.0177 |
| *O-15 with*<br>*washout* | 1042 | 10.37 | 0.01 | 0.00 | 0.00 | 0.00 | 0.00 | 0.00 | 0.00 |
| *C-11 with*<br>*washout* | 61.6724 | 20.9482 | 9.4747 | 3.0319 | 0.3105 | 0.0994 | 0.0318 | 0.0102 | 0.0033 |

### 5.2 PET imaging of low activity concentration

The low activity concentration measurements are presented in Figure 3. In (A) the image of a 3 mL syringe is shown on the left with an activity concentration of 1 kBq/mL and on the right with activity concentration of 11 Bq/mL. One can observe a lot of noise in the low activity concentration on the right. The noise is present also outside of the syringe.

**Figure 3.**
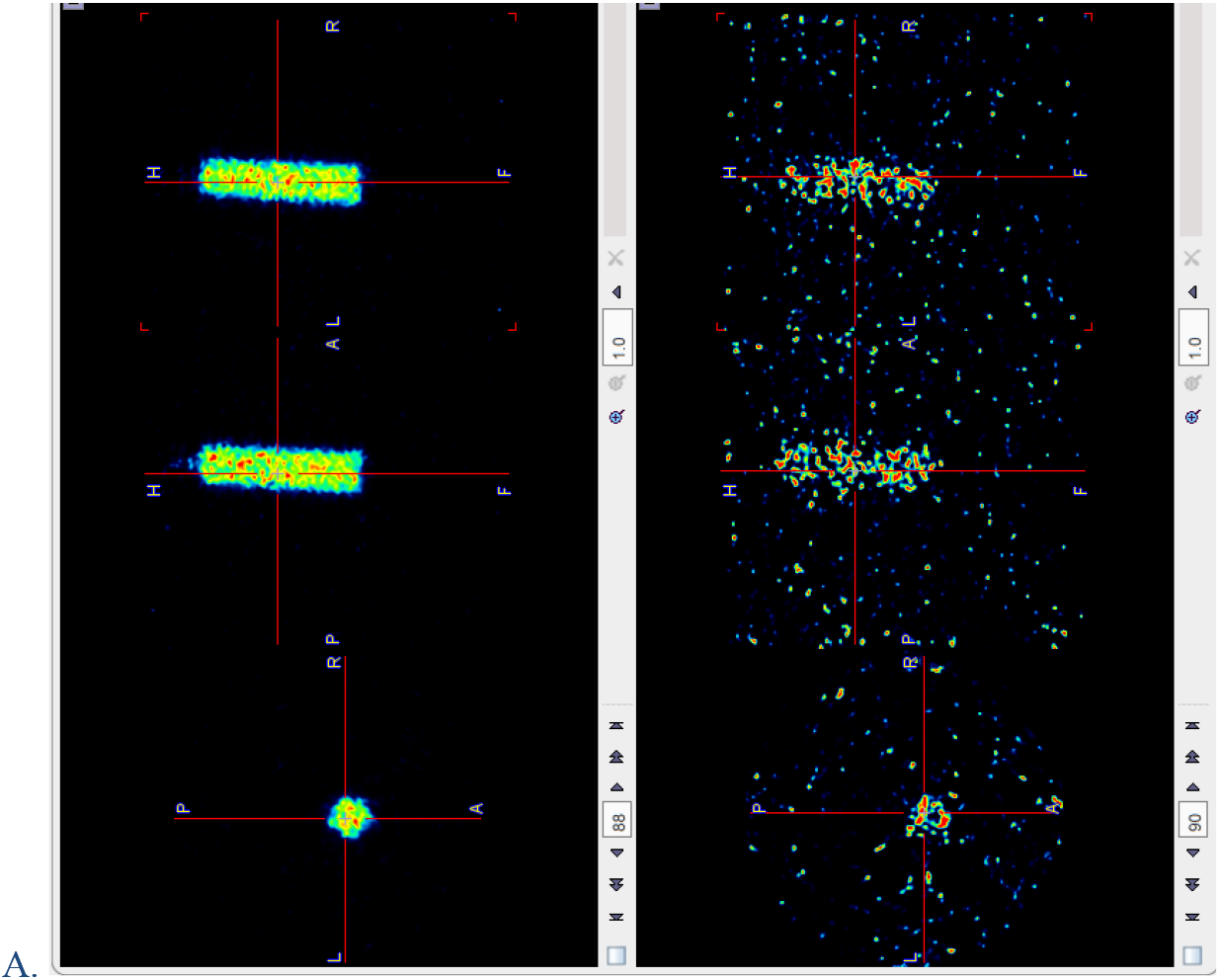
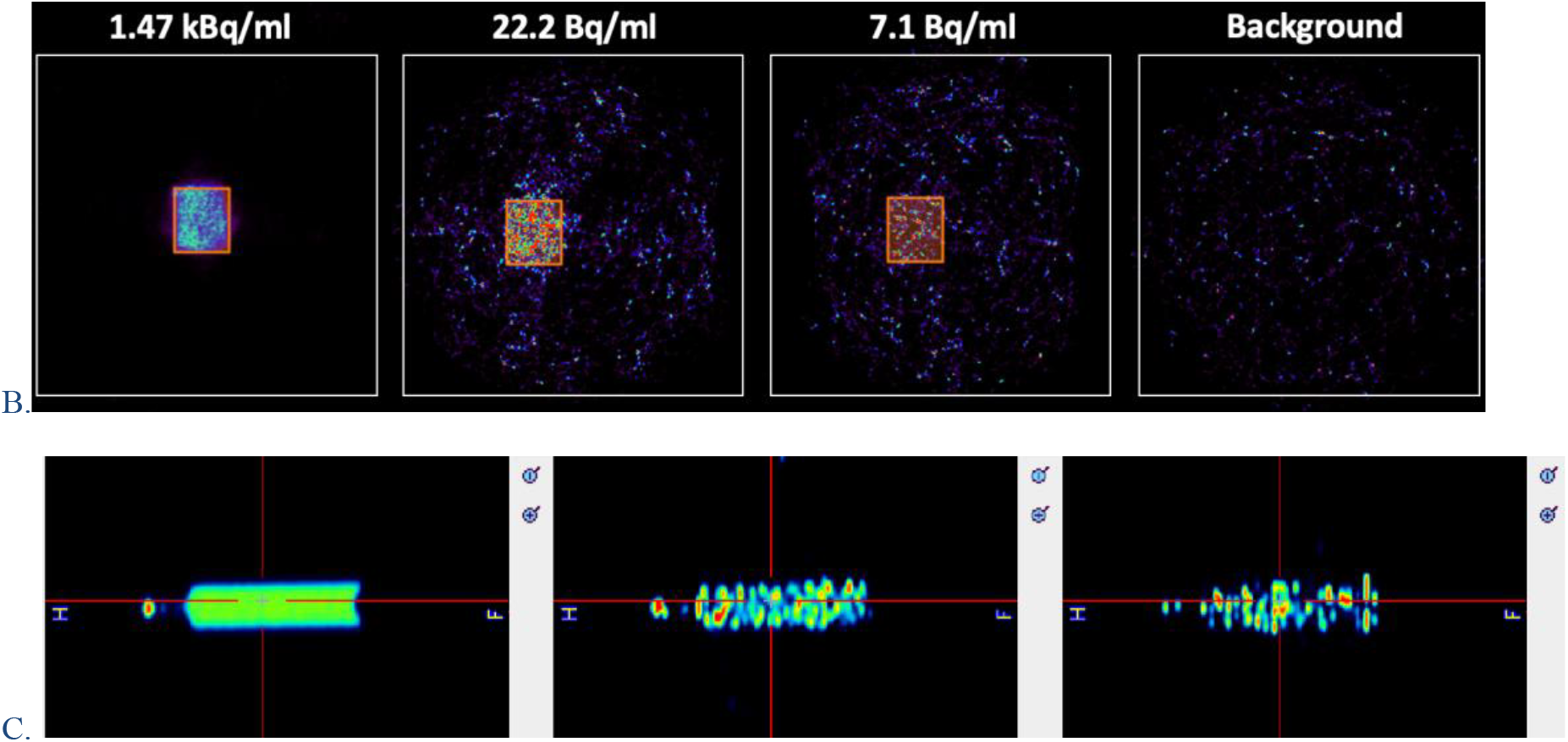
Low activity concentration measurement in PET. A. 3 mL syringe (Φ = 10 mm, H=40 mm) filled with [F-18]FDG. 60 min acquisition in Albira microPET scanner of 5% sensitivity. **Left**: at study start with 1 kBq/mL.**Right**: at 12 hours with 11 Bq/mL. Note random pixels outside of syringe due to Lu-176 PET LYSO crystals activity. B. Reconstructed images of the 20.25 mL vial at different concentration values and reconstructed image of the background. C. Activity measured in BGO-based scanner of a syringe of 30 mL of [F-18]fluorodeoxyglucose. **Left:** activity concentration of 1 kBq/mL, **center** 1 Bq/mL (corresponding to 30 Gy immediately after from treatment in brain tissue), **right** 0.3 Bq/mL (10 Gy).

To verify that the noise is not accidental or specific to a model of PET scanner, we have performed a similar scan with 20.25 mL vial shown in Figure 3 (B). In this case, we have increased the volume of the vial to match the typical size of a treated tumor. We started from 1.47 kBq/mL and reached 7.4 Bq/mL that approximately corresponds to acquisition at Figure 3 (A). In the rightmost image we show background 1h scan that clearly shows the noise in all imaged volume, albeit less intense than in Figure 3 (A). In the second scan the coincidence window was reduced from 5 ns to 3 ns, indicating that it is a type of random noise. With the reduced coincidence window we expected less random coincidences, which was observed, albeit not quantified. If the noise was some due to some electronics, heat etc., it would stay the same or change randomly. The number of counts is consistent with Lu-176 natural isotope activity, which is about 456 disintegrations per mL of the LSO crystal(23). Both scanners used LYSO crystal detectors. Therefore, we proceeded to a scan in a PET which doesn’t contain lutetium in the crystals to reduce the background noise.

100 Gy is the dose considered in some FLASH treatments, not yet in the usual treatment. The typical dose per session is about 2-3 Gy. Therefore, we need this method to be able to detect such low doses without excessive noise. We chose a clinical BGO-based scanner of high sensitivity of 4 %, similar to preclinical ones. The results are shown in Figure 3 (C). We have measured a 30 mL syringe in 1 hour time window. We started from 1 kBq/ml and registered 1 Bq/mL corresponding to 33 Gy and 0.3 Bq/mL corresponding to 10 Gy without any noise outside of the vial. These measurements show feasibility of detecting doses used in proton therapy session.

### 5.3 DVH in gamma and simulated tumor in proton therapy

The example of benefit of using F-18 compared to C-11 is shown in Figure 5 A. Dose-Volume Histogram (DVH) of F-18 (red dots) follows the dose curve from Figure 2 imperfect plan on (blue), while the one for C-11 (green dash) is deviating. In Figure 5 B the dose profile along the beam is shown in blue with dose recalculated from F-18 activity concentration from Figure 2 (orange). The relative error inside the tumor is 3.1%, and along all the beam is 3.9%. For comparison we show the counts profile for C-11 (gray).

## 6 Discussion

From Figure 4 (reproduced under CC BY license from (24)) we can see that the reproduction of dose in PET image is of a very low quality if all positron emitters are included in the analysis. While planned dose distribution is practically symmetrical (left to right), the PET image does not reflect this symmetry, nor permits quantification of the dose. The reconstruction quality does not allow the use of this tool either for range verification or for dose verification.

**Figure 4.**
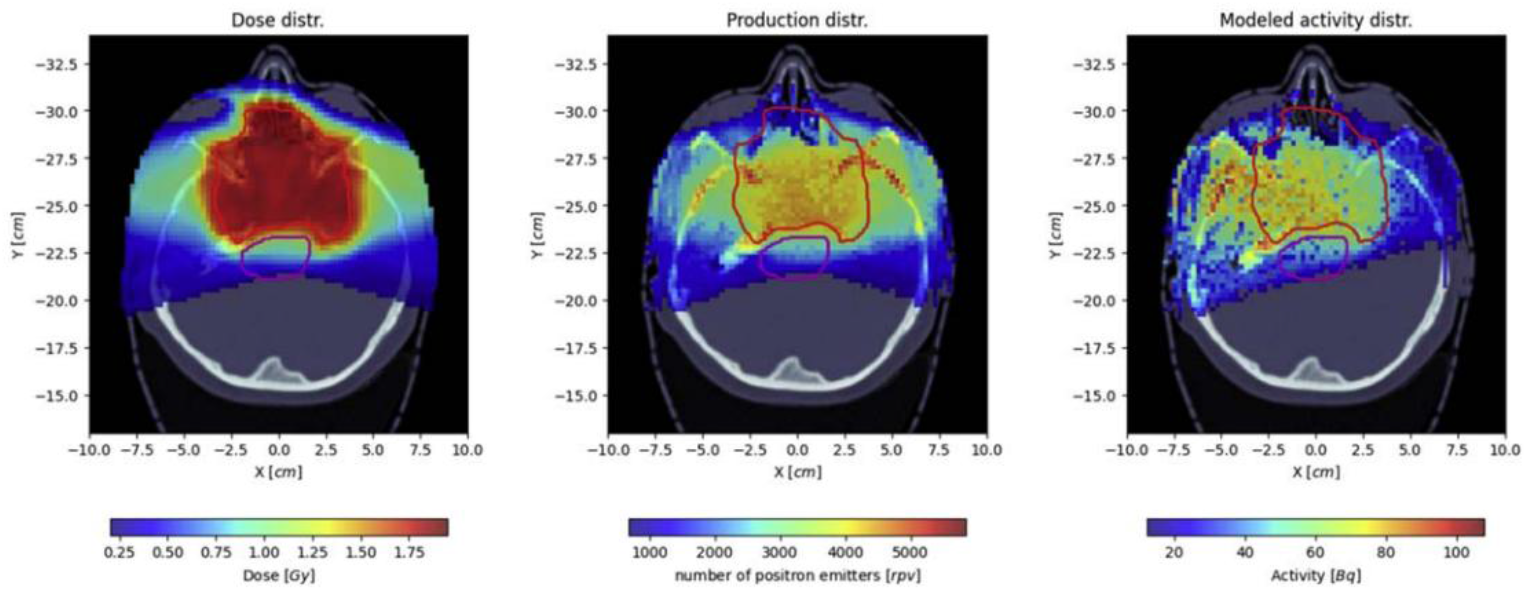
From reference (24) Left to right: Dose from treatment plan of 2 Gy (left), produced by all positron-emitting isotopes (per voxel of 2.5×2.5×2.5 mm, center) and modelled activity in Bq (supposedly per voxel, right), which scales 100 Bq/voxel to 6.4 kBq/mL. Reproduced according to CC BY license from Open Access article.

Figure 5 shows that F-18 distribution closely follows DVH of the planned dose. C-11 distribution deviates significantly from planned dose and therefore cannot be used as a dose map. The curves for N-13 and O-15 deviate still more dramatically than C-11 (graphs not shown).

**Figure 5.**
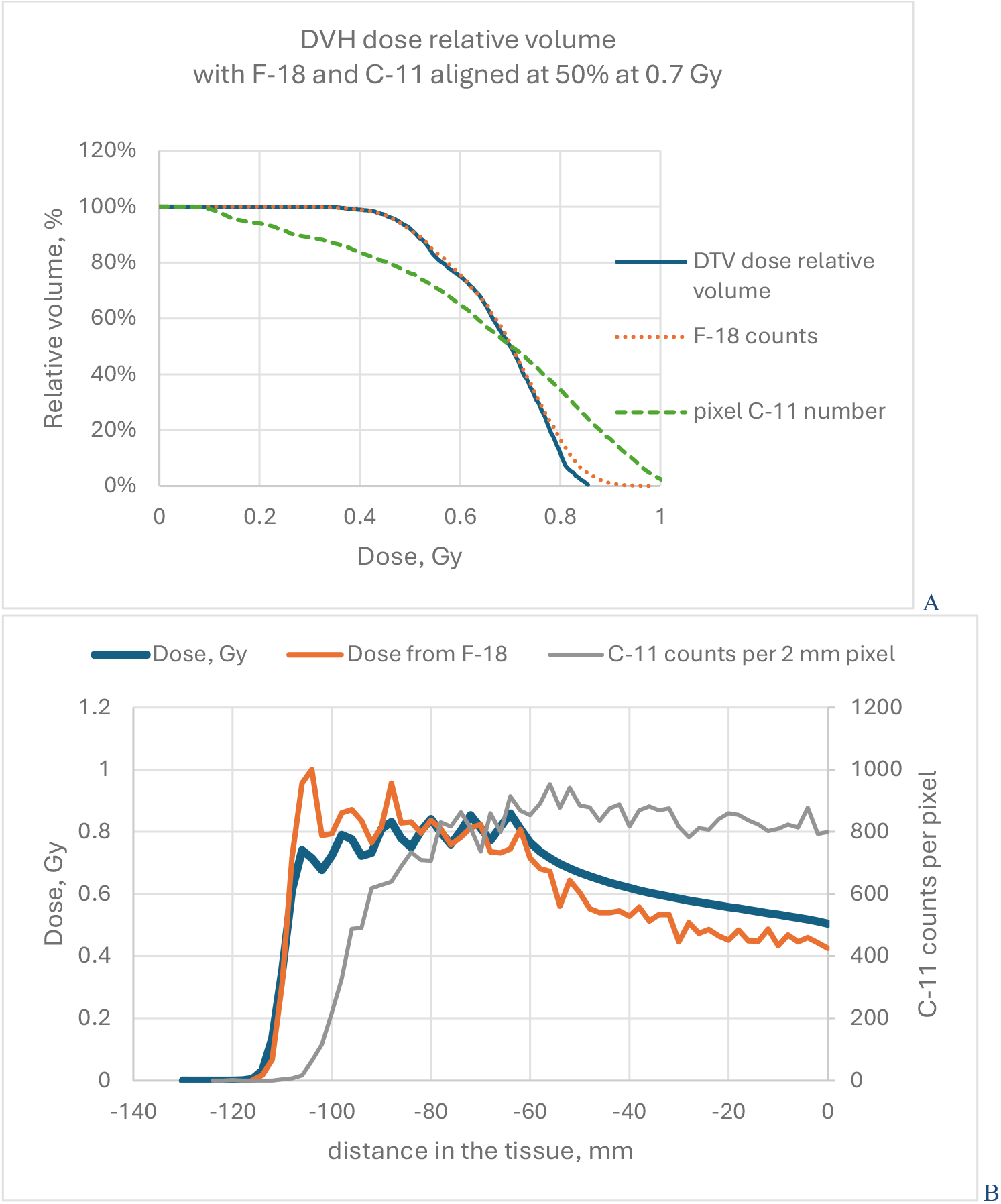
A. DVH analysis of dose for the plan shown in Figure 2 top. C-11 and F-18 isotope production in the simulated tumor volume in Figure 2 middle and bottom. B. Dose profile in the center of Figure 2 along beam direction (blue). Orange is a dose profile recalculated with factor for F-18 from Table 2 yielding 3.1% relative error in the tumor and 3.9% along all beam. Gray is the profile of C-11 counts along the same line (right vertical axis).

From Table 2 we can see that for the times below 240 min we may still see C-11 contribution, but the exact time will be determined in future experiments with small animal and human participation to determine washout.

The low activity concentration of F-18 produced immediately after proton irradiation is possible to measure with actual BGO based clinical scanners, however the sensitivity should be increased by higher solid angle coverage. The method is more suitable for high doses per session, these used in FLASH proton therapy.

### 6.1 Study limitations

The study has several limitations. The activity concentration measurements were done in phantoms with a known activity concentration and not on tissue-equivalent phantoms irradiated with proton. We did not determine if F-18 will stay in the production location and will not be metabolized or diffused. That needs to be determined in a small animal preclinical study or in patients within a clinical trial. The simulations of β+ nuclide distribution do not consider image degradation due to acquisition in a PET scanner.

### 6.2 Conclusion

This study demonstrates the feasibility of using F-18 PET imaging for in vivo dose verification in proton and helium-4 ion therapy. Simulations showed that F-18 production aligns with the planned dose distribution within 1–2 mm, significantly outperforming other positron-emitting isotopes such as C-11, N-13, and O-15. PET scanner measurements confirmed that clinical BGO-based systems can detect the low activity concentrations expected after therapeutic doses, without the background noise associated with lutetium-based detectors.

These findings suggest that F-18–based PET dosimetry offers a practical path forward for post-treatment dose verification in proton therapy, particularly for high-dose or FLASH applications. Future studies in preclinical models and clinical settings are needed to confirm isotope retention and to evaluate biological washout dynamics in vivo.

## Data Availability

All data produced in the present study are available upon reasonable request to the authors

## 7 Acknowledgements

The publication was funded in part by Fundación Vital Fundazioa.

We would like to thank Luiza Haberska for performing measurement of Figure 3 C.

## 9 Supplemental information

Supplemental Material 1 zip – macro of Gate 9.2

Supplemental Material 2 zip – macro of Gate 10.0.1

**Supplementary Figure 1.**
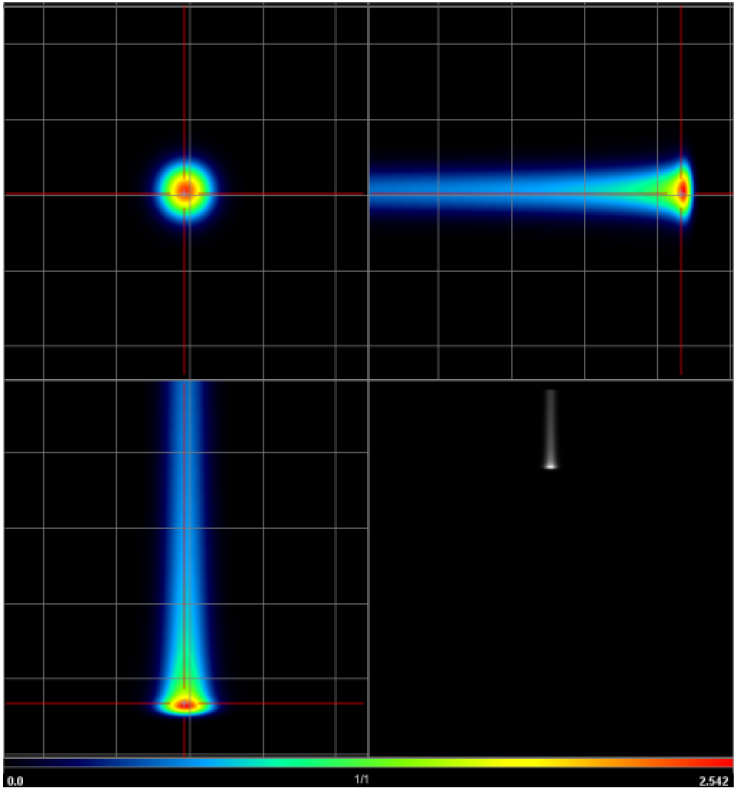
The simulated 3D dose profile of a 75 MeV needle proton beam in brain tissue. The full cold scale dose is 2.54 Gy at the tip of the red cross. The fourth quadrant is an MIP projection on the XZ plane. Note the widening of the beam close to the Bragg peak. The grid is 10 mm. The Bragg peak develops at about 43 mm depth.

**Supplementary Figure 2.**
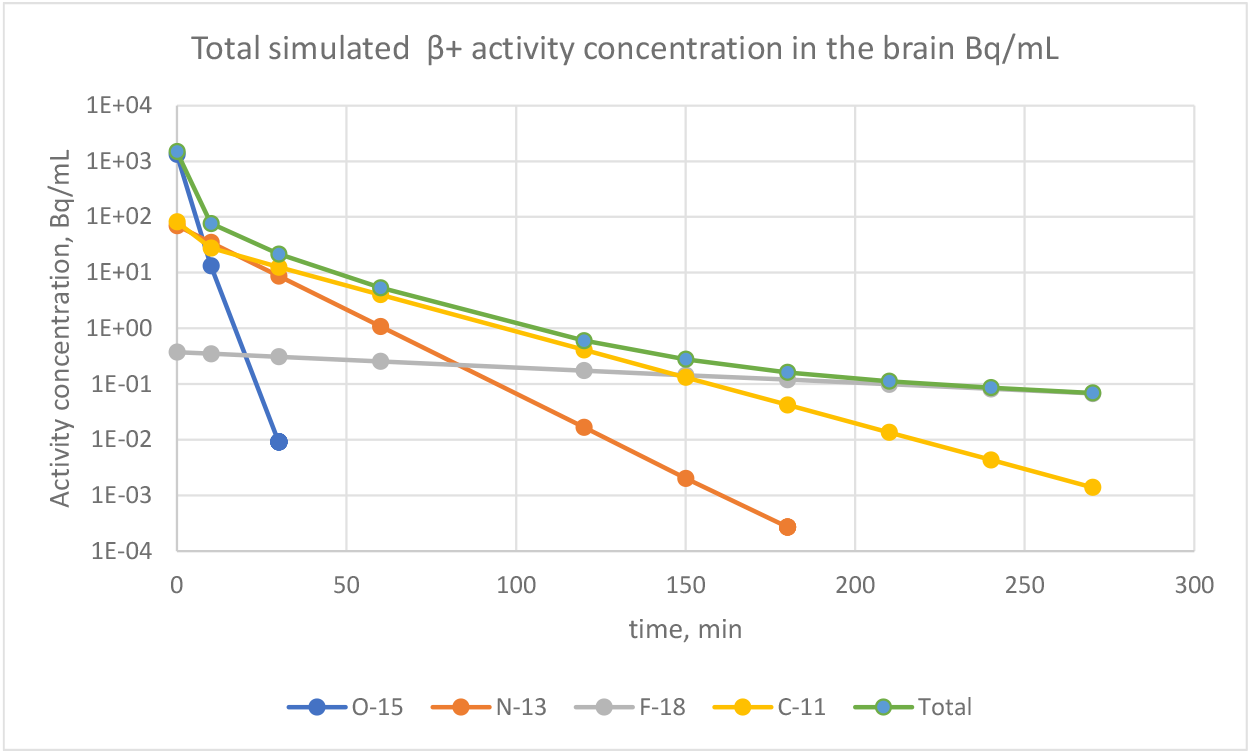
Brain phantom activity at 75 MeV p in needle beam in Suppl. Fig. 1.

**Supplementary Figure 3.**
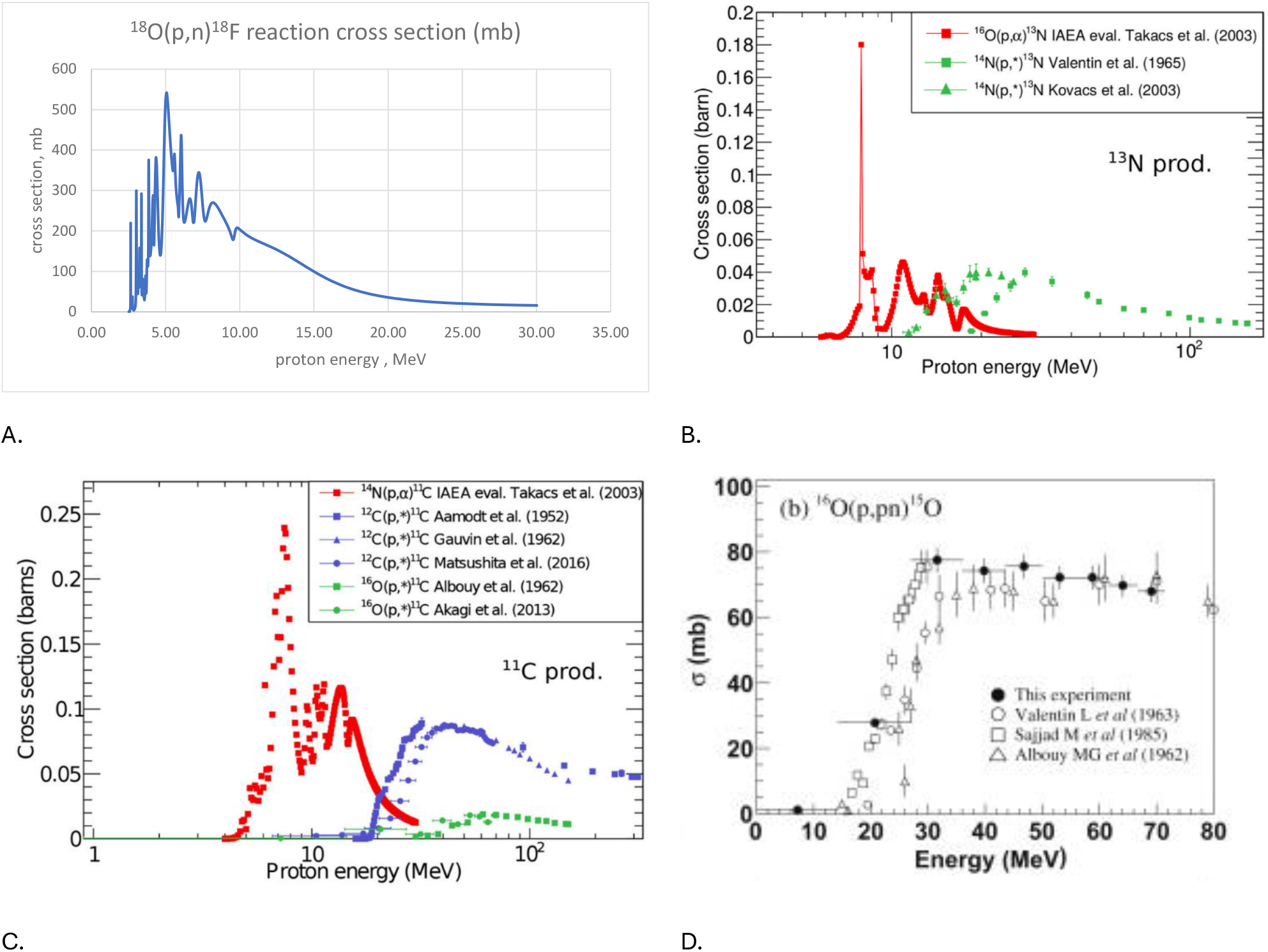
Cross Cross section vs. proton energy curves for four positron-emitting isotopes. A is the data from(13). B and C are taken from Reference(25)CC BY NC ND license, D from Reference(26), copied with permission. Note that A and D have horizontal linear energy scale, while B and C have logarithmic scale..

