## Supplemental Material 2 for "Precise dose verification in proton therapy using Positron Emission Tomography": RSplanBrain.py

```
#!/usr/bin/env python3
# -*- coding: utf-8 -*-
# If you use that code, please cite the relevant publication:
# "Precise dose verification in proton therapy using Positron Emission Tomography."
# Marcin Balcerzyk et.al. (2025) MedRxiv and subsequent peer-reviewd one. 
# to use this code you need Python 3.8 or higher and opengate version 10.0.1 with manually compiled branch isotopes or version 10.0.3 once it is available.

import matplotlib.pyplot as plt
import numpy as np
import opengate as gate
import pathlib
import pyvista
import multiprocessing
import SimpleITK as sitk
import opengate_core as gate_core
from user_colors import colors
from opengate.contrib.beamlines.ionbeamline import BeamlineModel
from opengate.contrib.tps.ionbeamtherapy import spots_info_from_txt


current_path = pathlib.Path(__file__).parent.resolve()
data_path = current_path / "data"
output_path = current_path / "output22"
# output_file = output_path / "dose.mhd"
# units
m = gate.g4_units.m
cm = gate.g4_units.cm
mm = gate.g4_units.mm
um = gate.g4_units.um
s = gate.g4_units.s
year = gate.g4_units.year
keV = gate.g4_units.keV
MeV = gate.g4_units.MeV
Bq = gate.g4_units.Bq # this is 1e-9 in utility.py

deg = gate.g4_units.deg

alpha_channel = -1


def simulation(visu,  x, y, z, n_sim):	# run_timing_intervals,


	# create the simulation
	sim = gate.Simulation()

	sim.progress_bar = True

	# main user options
	ui = sim.user_info
	ui.g4_verbose = False
	ui.g4_verbose_level = 2
	ui.visu = visu
	ui.visu_type = "vrml_file_only"
	ui.visu_filename = "visu_RSplanBrain.wrl" # f"visu_X_{x}_Z{z}.wrl"
	ui.random_seed = "auto"
	ui.number_of_threads = 10

	# materials
	sim.volume_manager.add_material_database(data_path / "GateMaterials.db")

	# change world size
	world = sim.world
	world.size = [500 * mm, 500 * mm, 500 * mm]
	world.material = "Air"
	fO15=sim.add_filter("ParticleFilter","fO15")
	fO15.particle = "O15"
	fC11=sim.add_filter("ParticleFilter","fC11")
	fC11.particle = "C11"
	fN12=sim.add_filter("ParticleFilter","fN12")
	fN12.particle = "N12"
	fN13=sim.add_filter("ParticleFilter","fN13")
	fN13.particle = "N13"
	fF18=sim.add_filter("ParticleFilter","fF18")
	fF18.particle = "F18"
	filterSet=[fO15, fC11, fN12, fN13, fF18]
	#filterSet = sim.add_filter("ParticleFilter", "filterSet")
	#filterSet.particle=["O15", "C11", "N12", "N13", "F18"]
	
	phantom, source = PhantomAndSource(x, y, z, sim, n_sim)
	# Brain tumor location that coincides with the range of protons and treatment plan.
	BrainTumor= sim.add_volume("Box", "BrainTumor")
	BrainTumor.size = [30 * mm, 30 * mm, 40 * mm] # 30x30x40 mm
	BrainTumor.material = "BrainO16_18"# "Brain"
	BrainTumor.color = colors.yellow
	BrainTumor.translation = [0, 0, 10 * mm] # 84 mm in Z direction, -10 mm from Brain/world center
	BrainTumor.mother = phantom.name

	#dose actor
	dose = sim.add_actor("DoseActor", "dose")
	dose.attached_to = phantom.name
	dose.output_filename = output_path / f"dose_to_{dose.attached_to}.mhd"
	dose.write_to_disk = True
	"""
	dose.dose_unit = "Gy"
	dose.dose_type = "DoseToWater"
	# dose.dose_type = "DoseToMaterial" # Dose to the material of the phantom
	dose.material = "Water"  # Dose to water
	dose.energy_deposition = True
	dose.energy_deposition_unit = "MeV/mm^3"  # Energy deposition in MeV/mm^3
	dose.energy_deposition_type = "EnergyDepositionToWater"  # Energy deposition to water
	"""
	dose.size=[50, 50, 75] # 140 pixels in each direction
	dose.spacing=[2*mm, 2*mm, 2*mm] # 1 mm spacing in each direction
	dose.hit_type="random"
	dose.dose.active=True
	dose.counts.active=True
	# add stat actor
	stats = sim.add_actor("SimulationStatisticsActor", "stats")
	stats.track_types_flag = True
	statsTumor = sim.add_actor("SimulationStatisticsActor", "statsTumor")
	statsTumor.track_types_flag = True
	statsTumor.attached_to = BrainTumor.name #statistics are taken from all World. It seems, that attaching is inefective.
	stats.output_filename = output_path / "stats.txt"
	stats.write_to_disk = True
	statsTumor.output_filename = output_path / "statsTumor.txt"
	statsTumor.write_to_disk = True


	# timing
	# sim.run_timing_intervals = run_timing_intervals # [[0, 10.0 * s]] # If you do not want to use source.n, use actity and this line
	# do for None, O15, N12, N13, C11, F18 
	add_dose_actor(sim, phantom, 2*mm, "O15", fO15)
	add_dose_actor(sim, BrainTumor, 2*mm, "O15", fO15)
	add_dose_actor(sim, phantom, 2*mm, "C11", fC11)
	add_dose_actor(sim, BrainTumor, 2*mm, "C11", fC11)
	add_dose_actor(sim, phantom, 2*mm, "N12", fN12)
	add_dose_actor(sim, BrainTumor, 2*mm, "N12", fN12)
	add_dose_actor(sim, phantom, 2*mm, "N13", fN13)
	add_dose_actor(sim, BrainTumor, 2*mm, "N13", fN13)
	add_dose_actor(sim, phantom, 2*mm, "F18", fF18)
	add_dose_actor(sim, BrainTumor, 2*mm, "F18", fF18)
	add_phase_space_actor(sim, phantom, "filterSet", filterSet)
	add_hits_collection_actor(sim, phantom, "filterSet",filterSet) #, None, None)
	add_phase_space_actor(sim, BrainTumor, "filterSet", filterSet)
	add_hits_collection_actor(sim, BrainTumor, "filterSet",filterSet)
	# add_phase_space_actor(sim, BrainTumor, "O15", fO15)
	#than add phase space actor for Brain 
	# start simulation
	sim.run()
	# Write simulation statistics to a plain text file
	with open(output_path / "stats.txt", "w") as f:
		f.write(stats.__str__())
	with open(output_path / "statsTumor.txt", "w") as f:
		f.write(statsTumor.__str__())
	# Since only one source is created, use its properties directly
	#total_activity = source.activity * ui.number_of_threads
	#total_volume = source.volume
	#print(f"Total volume of sources: {total_volume} mm^3")
	#print(f"activity concentration: {activity_concentration} Bq/mm^3")
	# print(f"Total source activity: {total_activity} Bq")

def add_production_and_stopping_actor(sim, volume, spacing, isotope, filter):
	actor = sim.add_actor("ProductionAndStoppingActor", f"prod_stop_{volume.name}_{isotope}")
	actor.attached_to = volume.name
	actor.user_info_defaults="stopping" #should be "production" (counts protons) or "stopping" (counts product and is defalult) or remove it
	# actor.isotope = isotope
	# actor.pixel_size = volume.spacing if hasattr(volume, "spacing") else None
	actor.size = [s / spacing for s in volume.size] #size is dimension in number of pixels for this actor.
	actor.spacing = [spacing, spacing, spacing] # if hasattr(volume, "spacing") else [2 * mm, 2 * mm, 2 * mm]
	actor.filters.append(filter) # filter for O15 and other particles
	actor.output_filename = output_path / f"prod_stop_{isotope}_{volume.name}.mhd"
	actor.write_to_disk = True
	return actor
	
def add_dose_actor(sim, volume, spacing, isotope, filter):
	actor = sim.add_actor("DoseActor", f"dose_{volume.name}_{isotope}")
	actor.attached_to = volume.name
	actor.size = [s / spacing for s in volume.size]
	actor.spacing = [spacing, spacing, spacing]
	actor.filters.append(filter)
	actor.output_filename = output_path / f"dose_{isotope}_{volume.name}.mhd"
	actor.write_to_disk = True
	actor.hit_type="random"
	actor.dose.active=True
	actor.counts.active=True
	return actor

def add_phase_space_actor(sim, volume, isotope, filter):
	actor = sim.add_actor("PhaseSpaceActor", f"phsp_{volume.name}_{isotope}")
	#print("phsp name:"+f"phsp_{volume.name}") 
	actor.attached_to = volume.name
	# actor.output_filename = output_path / f"phsp_{isotope}_{volume.name}.root"
	actor.output_filename = output_path / f"phsp_{isotope}_{volume.name}.root"
	actor.write_to_disk = True
	actor.store_absorbed_event = False #True
	actor.steps_to_store = "first"
	actor.filters_boolean_operator = "or" # "or" or "and"
	
	#print all possible attributes
	#am = gate_core.GateDigiAttributeManager.GetInstance()
	#print(am.GetAvailableDigiAttributeNames())
	
	for f in filter:  # filterset is a list of filters
		actor.filters.append(f)
	#actor.filters.append(filter) # filter for O15 and other particles
	#actor.filters.append(filter2)
	
	actor.attributes = [
        "RunID",
        "EventID",
        "TrackID",
        "TrackCreatorProcess",
        "KineticEnergy",
        "Position",
		"EventPosition",
		"EventKineticEnergy",
		"EventDirection",
        "Direction",
        "GlobalTime",
		"LocalTime",
		"ParticleName",
		"PDGCode",
		"PrePositionLocal",
		"PreDirectionLocal",
    ]
	return actor
def add_hits_collection_actor(sim, volume, isotope, filter):
	actor = sim.add_actor("DigitizerHitsCollectionActor", f"hits_{isotope}_{volume.name}")
	actor.attached_to = volume.name
	actor.output_filename = output_path / f"hits_{isotope}_{volume.name}.root"
	actor.write_to_disk = True
	actor.attributes = [
		"RunID",
		"EventID",
		"TrackID",
		"ThreadID",
		"TrackCreatorProcess",
		"KineticEnergy",
		"Position",
		"EventPosition",
		"EventKineticEnergy",
		"EventDirection",
		"Direction",
		"GlobalTime",
		"LocalTime",
		"ParticleName",
		"PDGCode",
		"PrePositionLocal",
		"PreDirectionLocal",
	]	
	for f in filter:  # filterset is a list of filters
		actor.filters.append(f)
	actor.filters_boolean_operator = "or" # "or" or "and"
	# print("filter operator for Hits: "+str(actor.filters_boolean_operator))
	#actor.filters.append(filter) # filter for O15 and other particles
	#print all possible actor attributes
	#am = gate_core.GateDigiAttributeManager.GetInstance()
	#print(am.GetAvailableDigiAttributeNames())
	return actor
def PhantomAndSource(x, y, z, sim, n_sim):
	# create the phantom
	# Create the main brain phantom as a sphere
	phantom = sim.add_volume("Box", "phantom") #("Sphere", "phantom")
	#phantom.rmin = 0 * mm
	#phantom.rmax = 70 * mm
	phantom.size = [100 * mm, 100 * mm, 150 * mm] # 100^2 x 150 mm cube as in Raystation Aire_1
	phantom.material = "BrainO16_18" #  "Brain" to use natural oxygen (and no need for isotope branch compiled), "BrainO16_18" for O16 and O18 and isotope branch compiled or gate version 10.0.3
	phantom.color = colors.blue
	phantom.translation = [x, y, z]
	
	sim.physics_manager.enable_decay = True
	# create the source
	# beamline model
	beamline = BeamlineModel()
	beamline.name = None
	beamline.radiation_types = "proton"
	beamline.distance_nozzle_iso = 250 * mm  # distance from nozzle to isocenter, starts at the edge of the world 

	# polinomial coefficients
	beamline.energy_mean_coeffs = [1, 0] # original one [-1259.5, 912726, 1.00e7] powers are  2,1,0
	beamline.energy_spread_coeffs =[0.01 ]# [0.4417036946562556]
	beamline.sigma_x_coeffs = [0.00016686, -0.076132159, 11.88305136]# 5.82 mm, value for 100 MeV to start with. Original was [2.3335754] #[0.00016686,-0.076132,11.883]
	beamline.theta_x_coeffs = [-2.6000E-06, 0.00050119, -0.018560476]# 0.00559rad original[2.3335754e-3] # at 100 MeV 0.00559
	beamline.epsilon_x_coeffs =[0.0241] # mm*rad divergence, Gate: emittance. Original  [0.00078728e-3]
	beamline.sigma_y_coeffs = [0.00016686, -0.076132159, 11.88305136] #5.82 mm,  original [1.96433431] #[0.00016686,-0.076132,11.883]
	beamline.theta_y_coeffs = [-2.6000E-06, 0.00050119, -0.018560476]# 0.00559 rad [0.00079118e-3]
	beamline.epsilon_y_coeffs = [0.0241] # mm*rad divergence, Gate: emittance. Original[0.00249161e-3]

	# tps
	#n_sim = 1e6  # particles to simulate per beam
	tps = sim.add_source("TreatmentPlanPBSource", "TPSource") # IonPencilBeamSource to get beam from Z direction, TreatmentPlanPBSource from XY direction. 
	# 0 deg is from -Y, 90 deg is from -X, 180 deg is from +Y, 270 deg is from +X 
	tps.n = n_sim
	tps.beam_model = beamline
	tps.plan_path = data_path / "PlanDescriptionToGATE3x3x3cm.txt" # This is  correct "PlanDescriptionToGATE3x3x3cm11Layers.txt" "TreatmentPlan4Gate-F5x5cm_E120MeVn.txt" #"Errors in text PlanDescriptionToGATE3x3x3cm.txt"
	tps.beam_nr = 1
	tps.gantry_rot_axis = "x"
	tps.sorted_spot_generation = False #if True, the source will be scanned spot by spot
	tps.particle = "proton"

	return phantom, beamline

def visualisation():
	pl = pyvista.Plotter()
	pl.import_vrml(f"visu_RSplanBrain.wrl") #"visu_X_{x}_Z{z}.wrl"
	pl.add_axes(line_width=5, color="white")
	pl.background_color = "black"
	for actor in pl.renderer.GetActors():
		actor.GetProperty().SetOpacity(.7)
	pl.show()

def run_simulation_in_process(activity_concentration, enable_visu, run_timing_intervals, xi, y, zi):
	simulation(activity_concentration, enable_visu, run_timing_intervals, xi, y, zi)

def main():
	#n = 1e-1	# See sim.run_timing_intervals - to have activity and duration correct
	activity_concentration = 1 # 1*0.031725235e-3 # *Bq/mm^3 , 0.031725235 Bq/microL/Gy corresponds to 3 Gy at start of the simulation 
	x, y, z = [0,0,0]
	# [0,5,10, 15, 25, 50, 75, 100], 0, [0,12.5] # position of the source in mm
	enable_visu = False # Set to True to enable visualisation
	# run_timing_intervals = [[0, 1.0 * s]] # 3600 s. If you do not want to use source.n, use actity and this line
	n_sim = 1e7 # particles to simulate per beam, this is the number of particles in the source
	""" processes = []
	for zi in z:
		for xi in x:
			p = multiprocessing.Process(
				target=run_simulation_in_process,
				args=(activity_concentration, enable_visu, run_timing_intervals, xi, y, zi)
			)
			p.start()
			p.join()  # Wait for each simulation to finish before starting the next
	"""
	simulation(enable_visu,  x, y, z, n_sim)
	if enable_visu:
		visualisation()


if __name__ == "__main__":
	main()
```
