## Supplemental Material 2 for "Precise dose verification in proton therapy using Positron Emission Tomography": user_colors.py

```
from box import Box

colors = Box({
	"invisible": [0, 0, 0, 0],
	"red": [1, 0, 0, 1],
	"green": [0, 1, 0, 1],
	"blue": [0, 0, 1, 1],
	"cyan": [0, 1, 1, 1],
	"magenta": [1, 0, 1, 1],
	"yellow": [1, 1, 0, 1],
	"gray": [.7, .7, .7, 1],
	"white": [1, 1, 1, 1],
	"pink": [1, 0.75, 0.79, 1],
	"orange": [1, 0.5, 0, 1],
})
```
